# Exploring the Association of Systolic Blood Pressure and Intracranial Pressure Variability and Subarachnoid Hemorrhage Patient Outcomes

**DOI:** 10.1101/2025.04.04.25324767

**Authors:** Saad Pirzada, Jane Quackenbush, Joshua Olexa, Abbey Kim, Yiting Lin, Stephanie Cardona, Quincy K Tran

## Abstract

**Background:** Subarachnoid hemorrhage (SAH) results from extravasation of blood into the subarachnoid space and is associated with high morbidity and mortality. This study aimed to compare systolic blood pressure variability (SBPV) and intracranial pressure variability (ICPV) in three 8-hour intervals during the first 24 hours after hospital admission and investigate their associations with discharge disposition and in-hospital mortality.

**Methods:** We retrospectively reviewed charts of adult patients with spontaneous, non-traumatic SAH admitted for at least 24 hours from 2016-2020. Hourly measurements were recorded for both systolic blood pressure (SBP) and intracranial pressure (ICP), and SBPV and ICPV were measured using successive variation (SV) and standard deviation (SD).

**Results:** A total of 240 patients were included (mean age 57±14.2 years, 64.6% female); 40 (16.7%) died. In the first 8-hour interval, higher SBP-SV (22.7±13.8) was significantly associated with mortality (p=0.028) and not being discharged home (p=0.022), compared to those who survived (17.6±7.5) or were discharged home (16.7±5.5). No significant differences in ICPV emerged for mortality or disposition, though higher ICP-SV in the first 8 hours approached significance (p=0.054) for discharge disposition. Receiver operating curve analysis showed poor discrimination for the first 8-hour SBP-SV (area under the curve 0.62) and failure for ICP-SV (0.51) in predicting mortality.

**Conclusions:** Greater SBP variability in the first 8 hours was linked to poorer outcomes, underscoring the potential importance of stabilizing blood pressure in this critical window. No clear association was observed between ICP variability and outcomes.

## Introduction

Subarachnoid hemorrhage (SAH) results from the extravasation of blood into the subarachnoid space. It is the third most common cerebrovascular disorder after acute ischemic stroke and intracerebral hemorrhage, and is associated with high morbidity and mortality.^1^ Most cases of SAH are traumatic; about 80% of spontaneous SAH result from aneurysmal rupture, which is associated with high rates of mortality.^1–3^ Recent research and clinical interest has been focused on the association of fluctuations of specific neurophysiologic measures such as blood pressure variability (BPV) and intracranial pressure variability (ICPV) and clinical outcomes in patients with SAH.^4,5^

For decades, aggressive management of high blood pressure (BP) has been foundational in the initial treatment of patients with SAH; current guidelines recommend lowering of systolic blood pressure (SBP) to equal or less than 140-160 mmHg (prior to securing of aneurysm).^1,6–8^ Accumulating evidence over more recent years suggests that BPV is also critical and adversely related to outcomes for this group of patients.^5,6,8,9^ Although the exact mechanism for why increased BPV worsens SAH patient outcomes is unknown, it is hypothesized that BPV may be a reflection of poor cerebral autoregulation.^4,9,10^

On the other hand, the scant literature on ICPV suggests that greater variation in short-term variations (55 to 15 second variations) in intracranial pressure (ICP) predicts better outcomes for patients with aneurysmal subarachnoid hemorrhage (aSAH) in the early post-ictal phase and early vasospasm phase, defined as days 1-3 and day 4-6.5, respectively.^4^ One possible explanation for this finding is that greater ICPV may indicate a healthier and more adaptive cerebrovascular system which can better regulate cerebral blood flow (CBF) and reduce the likelihood of secondary brain injury.^10^

Given the limited literature on systolic blood pressure variation (SBPV) and ICPV and its association on outcomes in patients with SAH, especially in time intervals that include the acute and hyperacute periods of SAH treatment, our study sought to compare SBP and ICPV in three separate 8-hour intervals during the first 24 hours of admission (the first, second, and third 8-hour intervals). We hypothesized that SBPV and ICPV would be greatest in the first 8 hours. Additionally, we explored both SBPV and ICPV and their association to outcomes in patients with SAH and compared the predictive power of these neurophysiologic measures. We hypothesized that greater SBPV is associated with worse patient outcomes and ICPV, given our variability timeframe of 8 hours and our outcome measures only encapsulating the first 24 hours, would not be associated with patient outcomes.

## Methods

This retrospective study included all adult patients (age ≥18) who experienced a non-traumatic, spontaneous SAH and were admitted to our institution for a minimum of 24 hours and monitored via an external ventricular drain (EVD). The study period was from 2016-2020.

Our study, “Exploring the Association of Systolic Blood Pressure and Intracranial Pressure Variability on Subarachnoid Hemorrhage Patient Outcomes”, was exempted by the Institutional Review Board at our institution (HP-00084554).

### Outcome Measures

The primary outcome of interest was in-hospital mortality. Secondary outcome of interest was discharge home, which was used as a surrogate for neurologic functional outcome. Prior studies have shown interrater discrepancies >23% when Cerebral Performance Category (CPC) scale or modified Rankin Scale (mRS) was used in a retrospective manner.^11–13^ Therefore, discharge location was used instead to assess for neurological functional outcome.

### Independent Variables

All patient data was obtained through the Electronic Medical Record (EMR). Some variables of interest included patients’ SBP, ICP, Glasgow Coma Scale (GCS), and Hunt and Hess scale grading (classification tool which helps assess severity and prognosis of patients with aSAH) (Table 1 and 2). At our institution, patients with SAH undergo BP and ICP measurements at least hourly for the first 24 hours. We sought to explore SBPV and ICPV through these hourly measurements, in three 8-hour intervals: the first 8 hours, the second 8 hours, and the third 8 hours. These variations can be calculated in many ways; for this study, we measured both SBPV and ICPV through successive variation (SV) and standard deviation (SD). The successive variation of systolic blood pressure (SBP-SV) is measured by taking the absolute difference between consecutive systolic blood pressure measurements and averaging these values. The standard deviation of systolic blood pressure (SBP-SD) is measured by taking the mean of the absolute difference of each systolic blood pressure measurement. The same applies for ICPV but using ICP instead of SBP measurements.

**Table 1.** Demographics and clinical characteristics of patients with SAH and between-group differences according to clinical outcomes.

|  | All Patients<br>(N=240) | Alive | Dead/<br>Hospice | Diff<br>erence | *P* | Not<br>dischar<br>ged<br>Home | Discharged<br>Home | Diff<br>erence | *P* |
| --- | --- | --- | --- | --- | --- | --- | --- | --- | --- |
| Age, years, mean<br>(SD) | 57 (14.16) | 57<br>(14) | 60<br>(14.9) | -3.4 | 0.188 | 59<br>(13.8) | 49 (12.2) | 10.4<br>7 | **0** |
| Gender, N (%) |  |  |  |  |  |  |  |  |  |
| Male | 85 (35.42) | 71<br>(35.5) | 14 (35) | 0.05 | 0.811 | 69<br>(37.3) | 16 (29) | 0.09 | 0.214 |
| Female | 155 (64.58) | 129<br>(64.5) | 26 (65) | -0.0<br>5 | 1 | 116<br>(62.7) | 39 (71) | -0.0<br>8 | 0.34 |
| Past medical<br>history |  |  |  |  |  |  |  |  |  |
| Hypertension,<br>N (%) | 155 (64.58) | 125<br>(63) | 30<br>(.75) | -0.1<br>3 | 0.102 | 124<br>(67) | 31 (56) | 0.11 | 0.157 |
| Diabetes, N<br>(%) | 42 (17.5) | 35<br>(18) | 7 (18) | 0 | 1 | 33 (18) | 9 (16) | 0.01 | 0.797 |
| Home<br>medications |  |  |  |  |  |  |  |  |  |
| Any<br>anticoagulation,<br>N (%) | 15 (6.25) | 12 (6) | 3 (7) | -0.0<br>2 | 0.738 | 13 (7) | 2 (4) | 0.04 | 0.281 |
| Any<br>anti-platelet, N<br>(%) | 60 (24.5) | 47<br>(24) | 13 (33) | -0.0<br>9 | 0.26 | 51 (28) | 9 (16) | 0.11 | 0.061 |
| Glasgow Coma<br>Scale |  |  |  |  |  |  |  |  |  |
| At admission,<br>median [IQR] | 9 [6-14] | 9<br>[6.25-14] | 5 [3-7] | 4 | **0** | 7<br>[5-13] | 13 [7-14] | -3 | **0** |
| At 24 hours,<br>median [IQR] | 9 [7-13] | 10<br>[8-14] | 5<br>[3.25-7<br>] | 4 | **0** | 8<br>[6-10] | 14 [10-15] | -4 | **0** |
| Admission<br>laboratory values |  |  |  |  |  |  |  |  |  |
| Sodium, mean<br>(SD) | 139.5 (4.3) | 139.3<br>4<br>(4.04) | 140.55<br>(5.38) | -1.2<br>1 | 0.184 | 139.76<br>(4.41) | 138.82<br>(3.88) | 0.94 | 0.13 |
| Platelet count,<br>mean (SD) | 234.9 (78.1) | 236.1<br>(66.8) | 229<br>(120) | 6.9 | 0.726 | 232<br>(81.7) | 245 (63.3) | -13 | 0.221 |
| Glucose, mean<br>(SD) | 168.47<br>(61.79) | 166.9<br>(62.3) | 176.1<br>(59.1) | -9.1 | 0.381 | 169.4<br>58.1) | 165.5 (73.3) | 3.9 | 0.719 |
| Lactate, mean<br>(SD) | 2.85 (5.95) | 2.77<br>(6.49) | 3.2<br>(1.83) | -0.4<br>3 | 0.486 | 3 (6.61) | 2.21 (0.96) | 0.79 | 0.162 |
| INR, mean<br>(SD) | 1.06 (0.13) | 1.051<br>(.124) | 1.103<br>(.142) | -0.0<br>5 | **0.04** | 1.073<br>(.136) | 1.0151 (.09) | 0.06 | **0** |
| Mechanical<br>ventilation, N (%) | 217 (90.42) | 177<br>(89) | 40<br>(100) | -0.1<br>1 | **0** | 180<br>(98) | 37 (67) | 0.31 | **0** |
| Any seizure during hospitalization, N (%) | 44 (18.33) | 34 (17) | 10 (25) | -0.08 | 0.276 | 37 (20) | 7 (13) | 0.07 | 0.176 |
| Craniectomy, N (%) | 101 (42.08) | 91 (46) | 10 (25) | 0.21 | **0.01** | 79 (43) | 22 (40) | 0.03 | 0.72 |
| Hunt Hess Scores, mean (SD) | 3.25 (1.16) | 3.07 (1.1) | 4.24 (.94) | -1.17 | **0** | 3.47 (1.15) | 2.61 (.92) | 0.86 | **0** |
| Vasospasm, N (%) | 137 (57.08) | 125 (62.5) | 12 (30) | 0.34 | **0** | 107 (60) | 30 (55) | 0.06 | 0.44 |

INR = international normalized ratio; IQR = interquartile range; SD = standard deviation.

**Table 2.** Fluid balance, in-hospital treatments, and neurophysiologic measures in patients with SAH and in-group differences according to clinical outcomes.

|  | All Patients<br>(N, %) | Alive | Dead/Hos<br>pice | *P* | Discharged<br>Home | Not<br>discharged<br>home | *P* |
| --- | --- | --- | --- | --- | --- | --- | --- |
| Balance of fluid in/out (SD) | 543 (1,976) | 451 (1,859) | 891<br>(2,355) | 0.225 | 263<br>(1,272) | 620 (2,127) | 0.131 |
| In-hospital treatment |  |  |  |  |  |  |  |
| Any anti-seizure medication, N (%) | 44 (18.3) | 34 (17) | 10 (25) | 0.332 | 7 (12.727) | 37 (20) | 0.3052 |
| Any hyperosmolar therapy, N (%) | 73 (30.4) | 49 (24.5) | 24 (60) | **<0.001** | 9 (16.36) | 64 (34.59) | **0.0158** |
| Blood products administration |  |  |  |  |  |  |  |
| None, N (%) | 178 (0.74) | 147 (0.735) | 31 (0.775) | 0.742 | 41<br>(0.7454) | 137 (0.741) | 1 |
| Any, N (%) | 61 (25.4) | 52 (26) | 9 (22.5) | 0.791 | 14 (25.45) | 47 (25.4) | 1 |
| Platelets, N (%) | 56 (0.233) | 48 (0.24) | 8 (0.20) | 0.733 | 13 (0.236) | 43 (0.232) | 1 |
| pRBCs, N (%) | 2 (0.0083) | 2 (0.01) | 0 (0) | 1 | 0 (0) | 2 (0.011) | 1 |
| FFP, N (%) | 3 (0.0125) | 2 (0.01) | 1 (0.025) | 1 | 1 (0.018) | 2 (0.011) | 1 |
| Neurophysiologic measures |  |  |  |  |  |  |  |
| *Blood Pressure Variability* |  |  |  |  |  |  |  |
| SBPmax, mean (SD) | 181.96<br>(26.93) | 180.54 (23.19) | 188.95<br>(40.37) | 0.209 | 180.76<br>(20.84) | 182.31<br>(28.51) | 0.661 |
| SBPmin, mean (SD) | 96.78 (20.66) | 96.99 (20.58) | 95.75<br>(21.30) | 0.737 | 97.04<br>(21.11) | 96.71<br>(20.58) | 0.919 |
| SBP-SD (first 8 hours), mean (SD) | 18.03 (7.24) | 17.49 (6.27) | 20.71<br>(10.57) | **0.069** | 17.16<br>(5.79) | 18.28 (7.6) | 0.245 |
| SBP-SD (second 8 hours), mean (SD) | 14.72 (6.65) | 14.37 (5.56) | 16.49<br>(10.43) | 0.217 | 14.23<br>(5.76) | 14.87 (6.89) | 0.497 |
| SBP-SD (third 8 hours), mean (SD) | 13.79 (7.44) | 13.6 (6.33) | 14.7<br>(11.680) | 0.579 | 12.68 (5.9) | 14.11 (7.82) | 0.152 |
| SBP-SV (first 8 hours) | 18.49 (9.03) | 17.64 (7.49) | 22.74<br>(13.84) | **0.028** | 16.65<br>(5.45) | 19.04 (9.80) | **0.022** |
| SBP-SV (second 8 hours) | 15.80 (9.07) | 15.57 (8.42) | 16.95<br>(11.83) | 0.48 | 15.05<br>(7.41) | 16.02 (9.5) | 0.433 |
| SBP-SV (third 8 hours) | 15.25 (7.6) | 15.26 (7.75) | 15.2<br>(7.07) | 0.96 | 14.66<br>(7.14) | 15.42 (7.78) | 0.51 |
| *Intracranial Pressure Variability* |  |  |  |  |  |  |  |
| ICP-SD (first 8 hours), mean (SD) | 3.68 (3.31) | 3.42 (2.28) | 5.00 (6.3) | 0.178 | 3.13 (1.94) | 3.84 (3.61) | 0.092 |
| ICP-SD (second 8 hours), mean (SD) | 3.06 (2.00) | 3.03 (1.82) | 3.2 (2.78) | 0.7 | 2.94 (1.90) | 3.09 (2.03) | 0.65 |
| ICP-SD (third 8 hours), mean (SD) | 3.05 (1.94) | 2.94 (1.76) | 3.64<br>(2.65) | 0.14 | 2.94 (1.86) | 3.08 (1.97) | 0.65 |
| ICP-SV (first 8 hours), mean (SD) | 4.19 (3.59) | 4.02 (3.08) | 5.04<br>(5.52) | 0.33 | 3.52 (2.01) | 4.39 (3.92) | **0.054** |
| ICP-SV (second 8 hours), mean (SD) | 3.64 (2.42) | 3.6 (2.24) | 3.82<br>(3.24) | 0.71 | 3.682<br>(2.44) | 3.63 (2.42) | 0.892 |
| ICP-SV (third 8 hours), mean (SD) | 3.67 (2.43) | 3.60 (2.33) | 4.05<br>(2.94) | 0.4 | 3.54 (2.21) | 3.71 (2.50) | 0.66 |

FFP = fresh frozen plasma; ICP = intracranial pressure; ICP-SD = standard deviation of intracranial pressure; ICP-SV = successive variation of intracranial pressure; pRBCs = packed red blood cells; SBP = systolic blood pressure; SBP-SD = standard deviation of systolic blood pressure; SBP-SV = successive variation of systolic blood pressure; SD = standard deviation.

### Data Collection and Management

All team members involved in data collection were trained by the principal investigator (PI). In the training process, all team members who collected data were first given a sample data collection of 10 patient charts and were required to achieve a minimum of 90% agreement with PI data before proceeding. All data was compiled into Microsoft Excel spreadsheet (Microsoft Corp, Seattle, WA). After data collection, a team member randomly checked 20% of the collected data to ensure an interrater agreement of at least 90% was maintained.

### Data Analysis

We employed descriptive analyses such as sample size (N) and percentage (%) for categorical variables and interquartile ranges (IQR), means, and SD for continuous variables.

We employed Mann-Whitney U test and independent sample T-test to compare continuous data between groups, including mortality (alive vs. dead) and discharge status (discharged home vs. not discharged home); Chi-Square test was used for categorical variables.

Given that SBP-SV was statistically significant in the deceased group, we sought to further examine the relation between SBP-SV in the first 8-hour interval and mortality by performing a univariate probit logit analysis. To further probe the discriminatory abilities of SBPV and ICPV in the varying 8-hour intervals for mortality and discharge home, an area under the receiver operating curve (AUROC) analysis was done. The AUROC analysis was parameterized for mortality such that 1 = dead and 0 = alive. For discharge home, it was such that 1 = not discharged home and 0 = discharged home. An AUROC <0.5 indicated no discriminatory ability, 0.5-0.6 was failed, 0.6-0.7 was poor, 0.7-0.8 was fair, 0.8-0.9 was good, and 0.9-1.0 was excellent.

Data analysis was conducted using both Minitab (Minitab Corp, State College, PA) and R version 4.4.2 (R Core Team, 2022).

## Results

Our study identified 240 patients with spontaneous, non-traumatic SAH admitted to our institution (Figure 1); 200 were aneurysmal and 40 were non-aneurysmal SAH. Of the 240, 155 were female (64.6%), and the mean age was 57 (± 14.2). The average age of patients discharged home was 49 (± 12.2), which was 10.5 years younger than those not discharged home (59 ± 13.8; p <0.001). Overall, a number of 40 patients died (16.7%) (Table 1). Upon admission, the median GCS score was 9 [IQR 6-14]. The admission GCS for patients who survived (9, [IQR 6.3-14]) was on average 4 points greater than the admission GCS of patients who did not survive (5, [IQR 3-7], p < 0.001). A total of 101 (42.1%) of the patients underwent a craniectomy, of these, 91 (46%) survived and 10 (25%) died (p = 0.01). The average Hunt and Hess score among all patients was 3.25 (±1.16). Patients who survived had a score of 3.07 (± 1.1) while those who died had a significantly higher score of 4.24 (± 0.94; p < 0.001).

**Figure 1.**
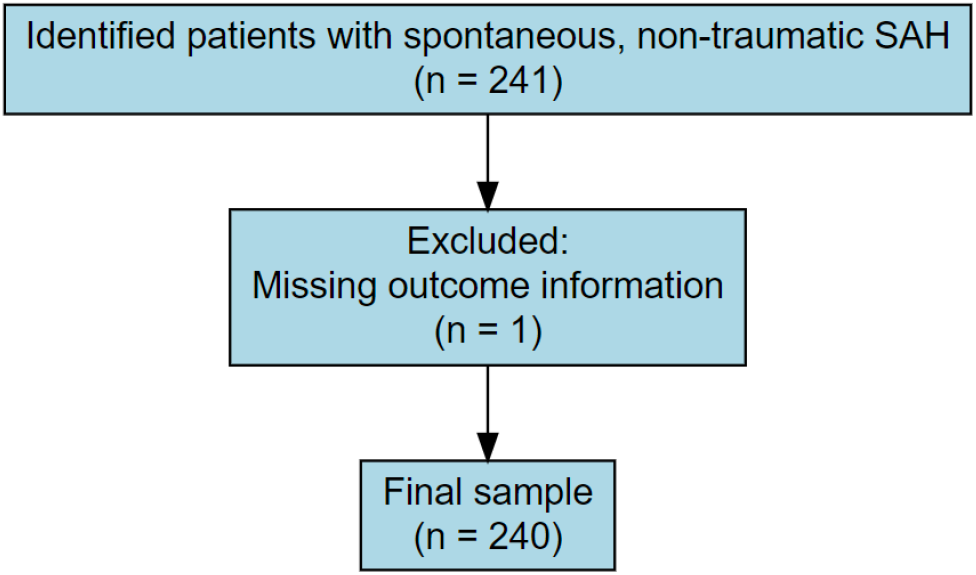
Flow Diagram of Patient Inclusion and Exclusion. SAH = subarachnoid hemorrhage.

Increased SBP-SV among SAH patients within the first eight hours of admission (first interval) was significantly greater in both patients who died and were not discharged home. Specifically, SBP-SV for patients who died was 22.7 (± 13.8), which was 5.1 points greater than those who survived (17.6 ± 7.5, p = 0.028). Among those not discharged home, the mean SBP-SV of 19.0 (± 9.8) was 2.4 points greater than those discharged home (16.7 ± 5.5, p = 0.022).

No intervals for ICPV were significantly different regarding mortality or disposition. However, ICP-SV in the first eight hours approached a significant difference in patients who were not discharged home (4.4 ± 3.9) compared to those discharged home (3.5 ± 2.0; p = 0.054).

### Probit Analysis: Mortality

Probit analysis was used to investigate the probability of mortality through a clinical predictor, specifically the continuous variable of SBP-SV. The univariate probit analysis found that SBP-SV, in the first eight hours, significantly predicted the outcome of mortality (p < 0.01). Furthermore, the results of our model suggest an SBP-SV of approximately 19 was associated with mortality in around 19% of our patient population (Figure 2).

**Figure 2.**
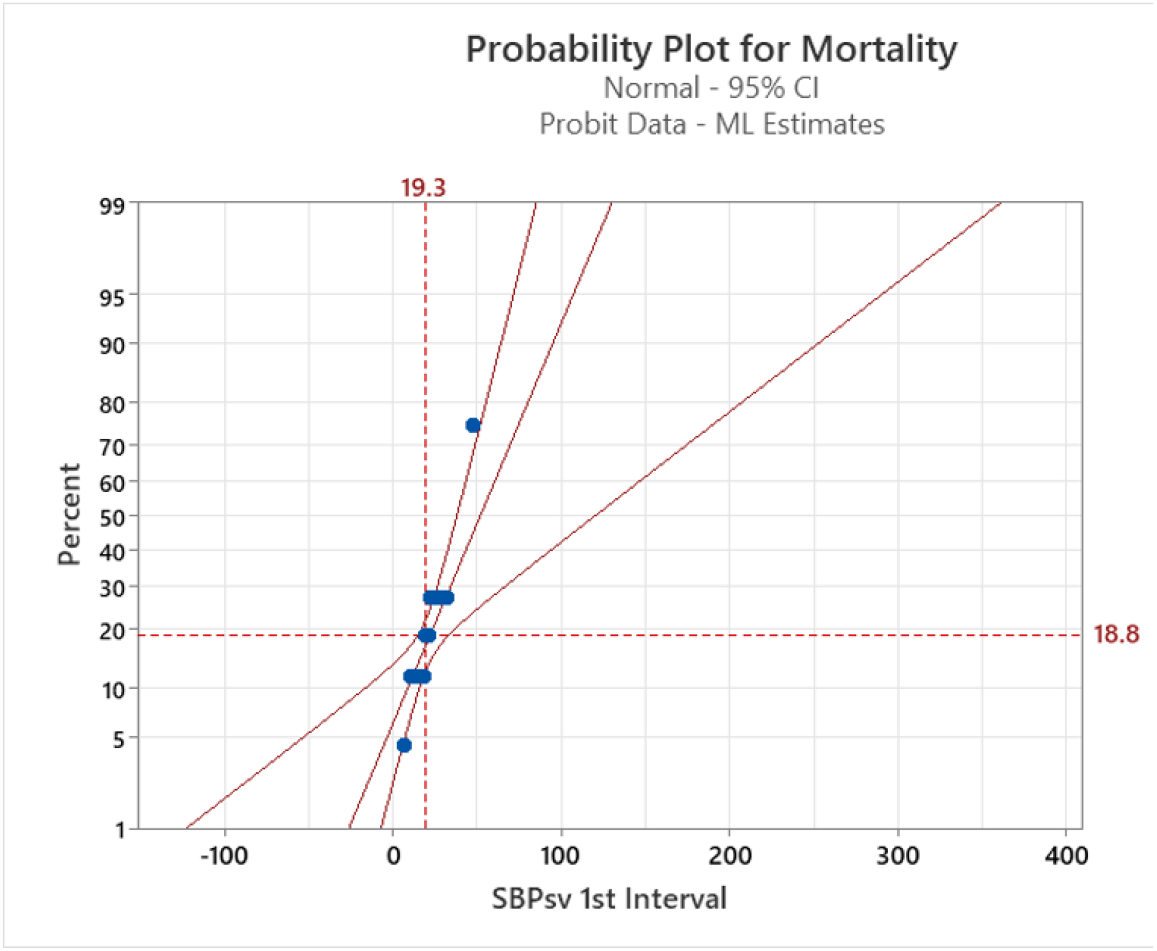
Probit Analysis – 1^st^ 8 hour interval. CI = confidence interval; ML = maximum likelihood; SBPsv = successive variation of systolic blood pressure.

### Receiving Operating Curve (ROC) Analysis: Mortality and Discharge Home

In the ROC used to predict mortality in the first interval, according to the standard interpretations of ROC curves, ICP-SV, with an area under the curve (AUC) of 0.51, failed to predict mortality whereas SBP-SV, with an AUC of 0.62, demonstrated poor discrimination (Figure 3).^14^ All other time intervals in regards to mortality and discharge home for both SBP-SV and ICP-SV failed at predicting outcomes (Figure 4; Supplemental Figure 1; Supplemental Figure 2).

**Figure 3.**
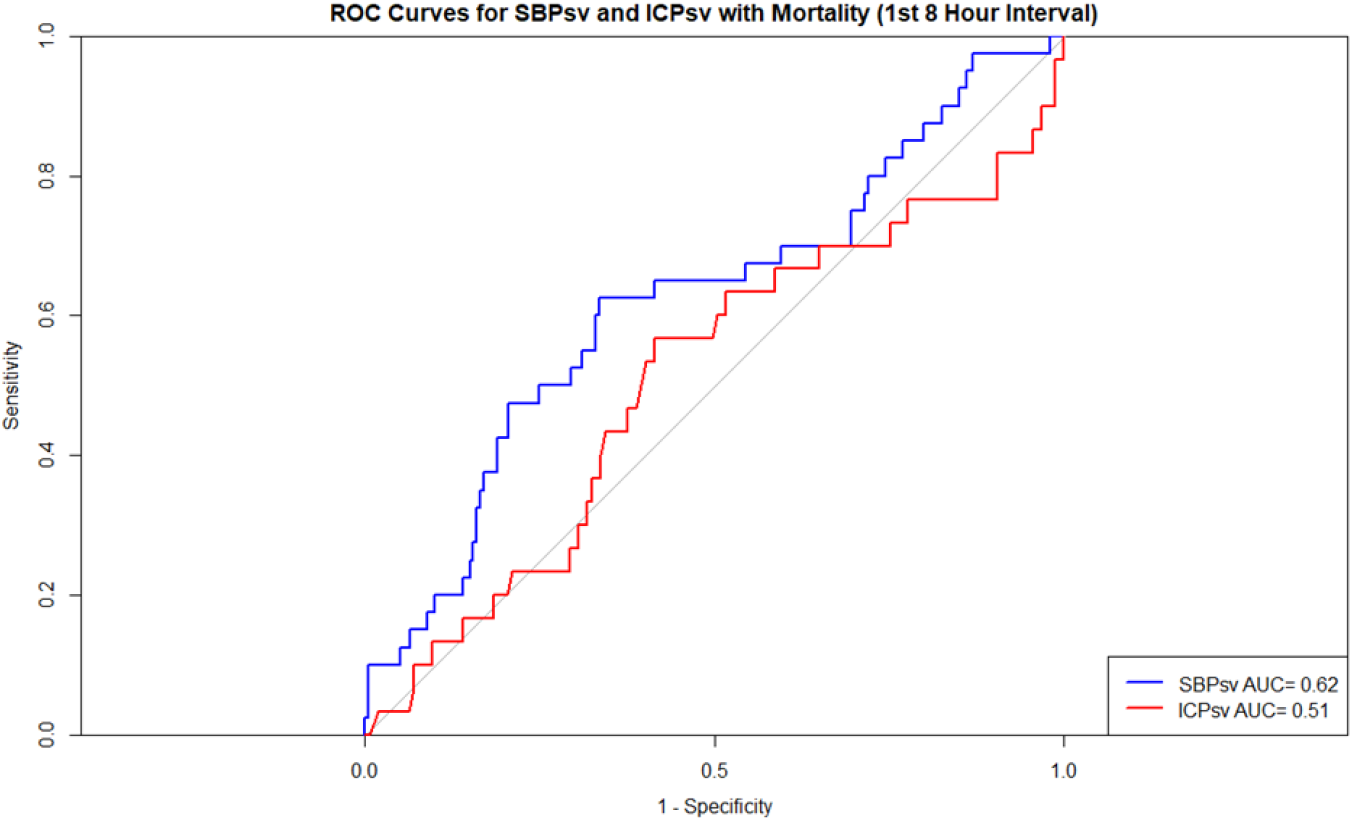
ROC Curves for Mortality - 1^st^ 8 hour interval. AUC = area under the curve; ICPsv = successive variation of intracranial pressure; ROC = receiver operating characteristic; SBPsv = successive variation of systolic blood pressure.

**Figure 4.**
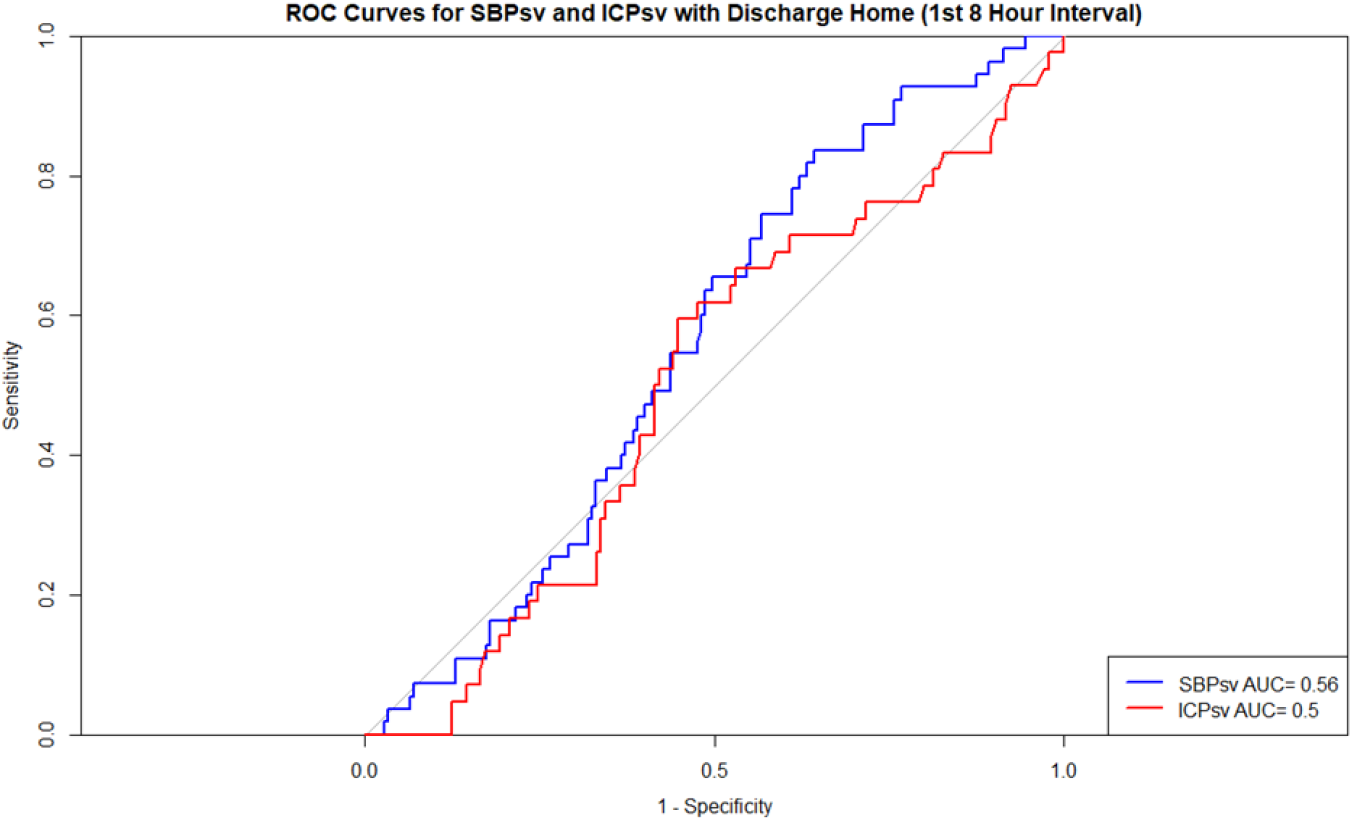
ROC Curves for Discharge Home - 1^st^ 8 hour interval. AUC = area under the curve; ICPsv = successive variation of intracranial pressure; ROC = receiver operating characteristic; SBPsv = successive variation of systolic blood pressure.

## Discussion

Previous research suggests greater ICPV in short-term variations, specifically variations between 55 to 15 seconds, is associated with decreased risk of cerebral delayed ischemia in aSAH patients in the early post ictal phase and early vasospasm phase.^4^ However, these associations in the early phases did not emerge when longer ICP variations were utilized, such as 30-minutes and 4-hours. Instead, associations between ICPV in 30-minutes and 4-hour parameters emerged in the late vasospasm phase (days 6.5-10). The short-term fluctuation in ICPV and its association with better outcomes is hypothesized to arise from the possible indication of healthier cerebral vessels with less atherosclerosis and cerebral vasospasm.^15^ Although the literature suggests higher short-term ICPV is associated with better clinical outcomes in aSAH patients in the early post-ictal and early vasospasm phase, our results showed no significant difference in ICPV in any of the time intervals for either mortality or discharge home. One possibility for the difference in results is that our ICPV was parameterized in 8-hour intervals, longer than any of the time intervals of the aforementioned study, where the longest parameter was 4-hours. Moreover, our outcomes of interest were mortality and discharge home and were not divided into different phases, such as in the study conducted by Wettervik et al., with phases of 1-3 days, 3-6.5 days, and 6.5-10 days.^4,10^ Differences in ICPV variability parameters and differences in outcome times do not allow for direct comparison to the current literature. However, our results suggest ICPV was reaching significance in the first interval (p = 0.054), with a greater ICP-SV in the first 8 hour interval seen in patients discharged home.

In our cohort of patients, SBP-SV in the first 8-hour interval was a significant BPV parameter associated with patient outcomes. As mentioned earlier, higher blood pressure variability in the form of SBP-SV in the first 8 hours of admission, was seen in patients who did not survive. This finding suggests that early stability in blood pressure variability is associated with better outcomes in SAH patients, similar to the findings of Cai et al.^16^ Cai et al. reported that higher variability in SBP-SV during the first 24 hours after endovascular coiling in poor grade aSAH patients, was a significant predictor of mortality and unfavorable neurologic outcome at 6 months. Our findings shed additional nuances to the work of Cai et al., as our study determined that SBP-SV was only significantly associated with mortality in the first 8-hour interval. One possible explanation for this finding is that a major early complication in aSAH patients is the occurrence of rebleeding, which occurs a majority of times (83%) within 12 hours after initial hemorrhage, with a median time of 180 minutes.^17^ Rebleeding following an aSAH is a significant predictor of mortality, with reported rates up to 60%.^18^ Moreover, the literature suggests that blood pressure variability is highly predictive of rebleeding in aSAH patients.^9^ Ultimately, we posit that less blood pressure variability in the first 8 hour interval may confer a protective effect, reducing the risk of rebleeding during an especially critical period and limiting the likelihood of a secondary brain injury.

Greater SBPV has been associated with a range of pathophysiologies, including myocardial infarction, stroke, and cerebral white matter lesions.^19,20^ However, beyond these associations, emerging evidence suggests that the time intervals from which SBPV occurs is critical to evaluating its prognostic value. Prior research from Kirkness et al. explored how the variability of blood pressure at different timescales was associated with 6-month outcomes in aSAH patients.^21^ Specifically, greater variability in short timeframes (5 seconds) was associated with greater survival after 6 months; however, greater variability in longer time frames (24 hours) was associated with decreased survival after 6 months. The authors suggest that the 24-hour slow variability reflects poorer adaptive mechanisms in response to physiological instability. Moreover, the 5-second fast variability is hypothesized to reflect a more responsive autonomic and cerebrovascular regulation. Our timeframe that was significantly associated with survival was the first 8-hour interval, which aligns more closely with the longer variability timeframe proposed by Kirkness et al. The findings of our study, further support the findings of Kirkness et al., that increased SBPV for longer periods of time is associated with decreased survival. Additionally, our work extends this hypothesis by positing that longer SBPV in the first 8-hours is significant in predicting survival whereas later time intervals (beyond the first 8 hours) are not.

In our cohort, SBP-SV in the first interval was significantly greater in patients who died (22.74) compared to those who survived (17.64; p = 0.028). As mentioned earlier, this difference may be a result of the early time window reflecting a critical period where blood pressure dynamics has the largest impact on survival. Specifically, high blood pressure variability, especially early on increases the likelihood of complications such as rebleeding, a significant predictor of mortality. Moreover, as described by Kirkness et al., large swings in blood pressure variation over larger periods of time may reflect poor adaptability to physiological instability and possibly explains the higher odds of mortality.

## Limitations

Our results highlight the significance of controlling SBPV during the early stages of treatment for SAH patients. However, this study also has multiple limitations that need to be accounted for. For instance, all of the data used in this study was collected retrospectively at a single institution. Our cohort and the management practices of our institution may not reflect that of other institutions. Additionally, SBP and ICP data was collected hourly over 24 hours, with the variability smoothed over 8-hour intervals. Prior work suggests that short-term fluctuations in blood pressure contain information that these longer intervals are unable to capture. It’s possible that the addition of these shorter time intervals would provide us with additional insights. Moreover, our ICPV was parameterized at 8-hour intervals, making it difficult to compare our findings with the literature with shorter time intervals. Furthermore, our results do not include long term neurological or functional status outcomes such as a 6-month GOS or modified Rankin Scale. Lastly, although the average SBP-SV in patients who died was higher, the overlap in individual values between those who survived and died makes it difficult to effectively discriminate between the two groups, explaining the AUC of 0.62, indicating poor discriminative ability.^22^ Moreover, the ROC curve seeks to find an optimal threshold value to separate mortality outcomes effectively. Still, with a large overlap in SBP-SV values, there is no significant threshold value, resulting in poor discrimination even if there are significantly different values between those who survived and died.^23^

## Conclusion

Our study determined that greater SBP variability in the first 8-hour interval was associated with poorer mortality outcomes, although with poor discriminatory ability (AUC = 0.62). When deciding management of SBPV, it is crucial for clinicians to evaluate the different clinical factors that predict outcomes and perform individual risk assessments.

## Data Availability

All data produced in the present study are available upon reasonable request to the authors

---

**Supplemental Figure 1.**
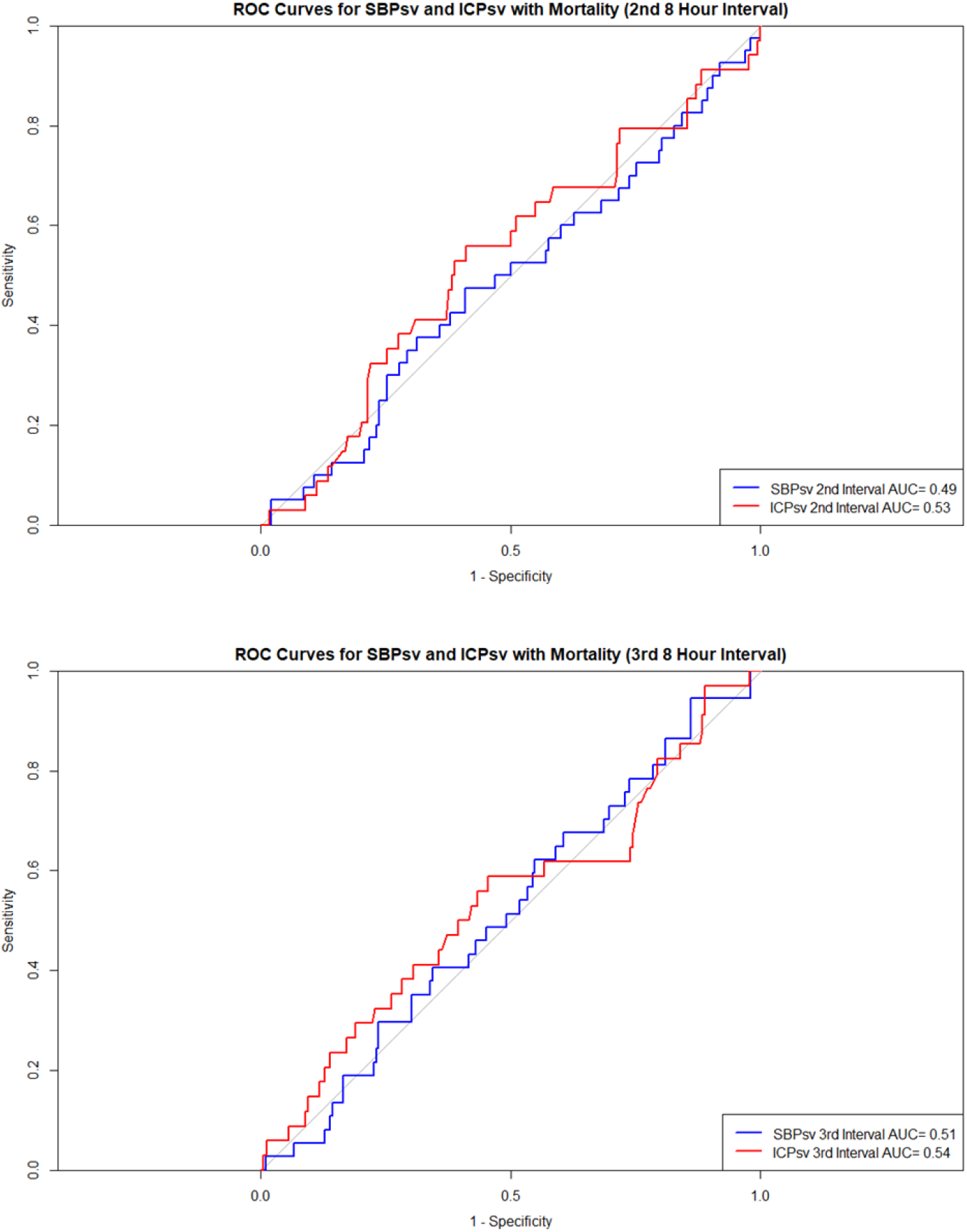
ROC Curves for Mortality 2^nd^ and 3^rd^ 8 hour interval. AUC = area under the curve; ICPsv = successive variation of intracranial pressure; ROC = receiver operating characteristic; SBPsv = successive variation of systolic blood pressure.

**Supplemental Figure 2.**
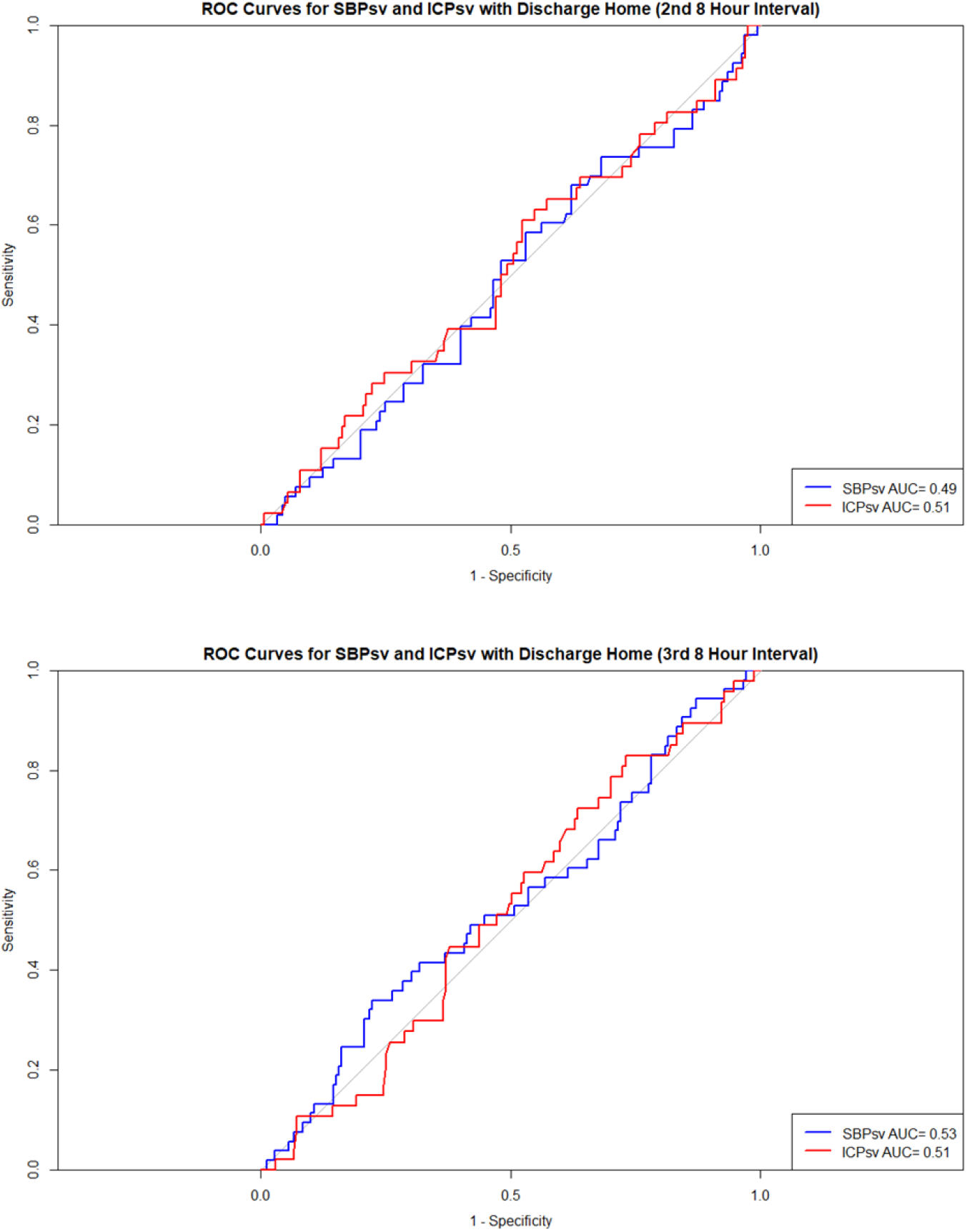
ROC Curves for Discharge Home 2^nd^ and 3^rd^ 8 hour interval. AUC = area under the curve; ICPsv = successive variation of intracranial pressure; ROC = receiver operating characteristic; SBPsv = successive variation of systolic blood pressure.

**Supplemental Table 1.**
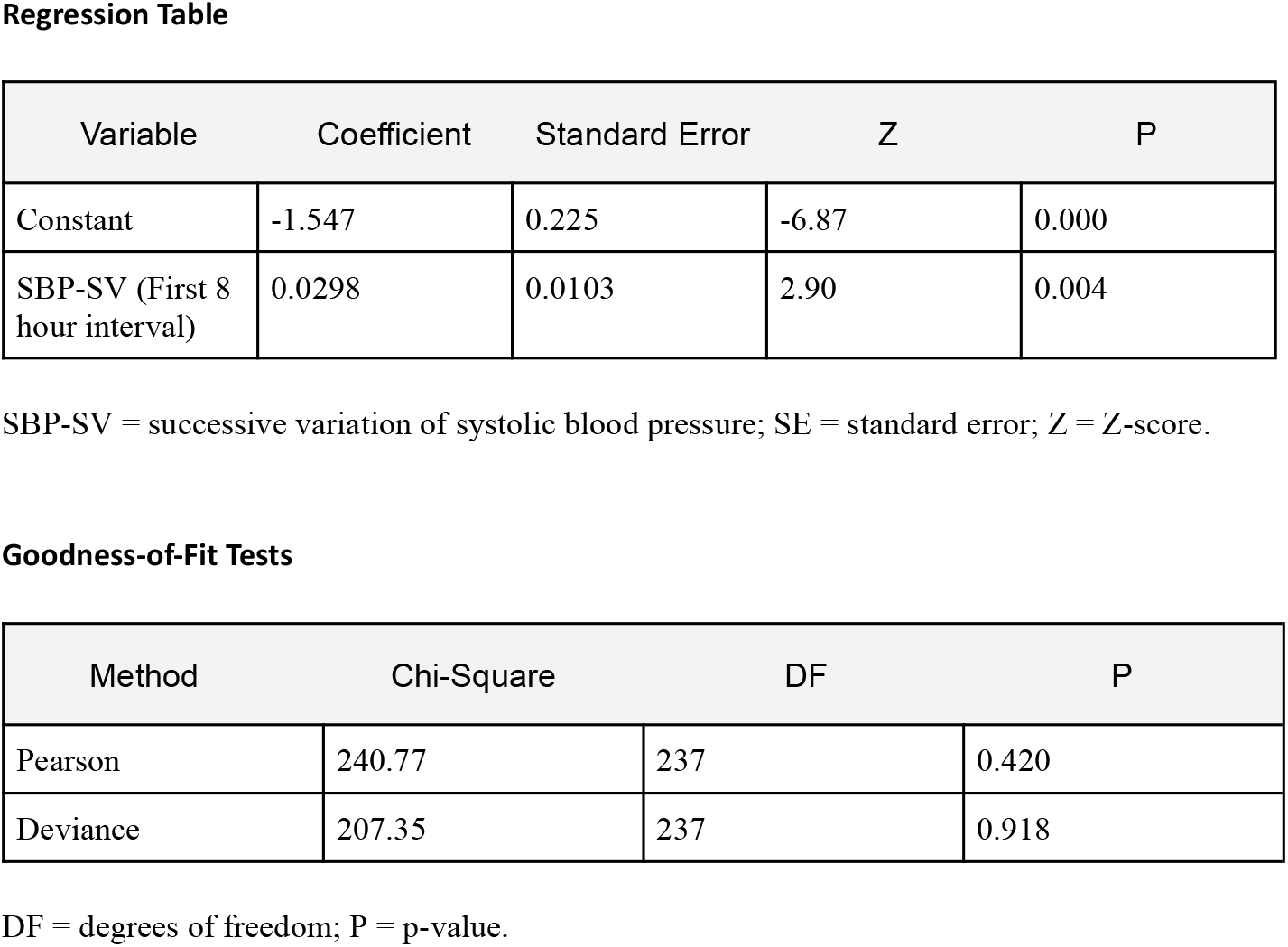
Regression Table and Goodness-of-Fit Tests.

## References

1. Darsie, Marin, Moheet, Asma, Lau, Winnie. The Pocket Guide to Neurocritical Care: A Concise Reference for the Evaluation and Management of Neurologic Emergencies. Neurocritical Care Society; 2019.

2. Stienen MN, Germans M, Burkhardt JK, et al. Predictors of In-Hospital Death After Aneurysmal Subarachnoid Hemorrhage: Analysis of a Nationwide Database (Swiss SOS [Swiss Study on Aneurysmal Subarachnoid Hemorrhage]). Stroke. 2018;49(2):333–340. doi:10.1161/STROKEAHA.117.019328

3. Van Gijn J, Rinkel GJE. Subarachnoid haemorrhage: diagnosis, causes and management. Brain. 2001;124(2):249–278. doi:10.1093/brain/124.2.249

4. Svedung Wettervik T, Howells T, Hånell A, Ronne-Engström E, Lewén A, Enblad P. Low intracranial pressure variability is associated with delayed cerebral ischemia and unfavorable outcome in aneurysmal subarachnoid hemorrhage. J Clin Monit Comput. 2022;36(2):569–578. doi:10.1007/s10877-021-00688-y

5. Yang M, Pan X, Liang Z, et al. Association between blood pressure variability and the short-term outcome in patients with acute spontaneous subarachnoid hemorrhage. Hypertens Res. 2019;42(11):1701–1707. doi:10.1038/s41440-019-0274-y

6. Connolly ES, Rabinstein AA, Carhuapoma JR, et al. Guidelines for the Management of Aneurysmal Subarachnoid Hemorrhage: A Guideline for Healthcare Professionals From the American Heart Association/American Stroke Association. Stroke. 2012;43(6):1711–1737. doi:10.1161/STR.0b013e3182587839

7. Diringer MN, Bleck TP, Claude Hemphill J, et al. Critical Care Management of Patients Following Aneurysmal Subarachnoid Hemorrhage: Recommendations from the Neurocritical Care Society’s Multidisciplinary Consensus Conference. Neurocrit Care. 2011;15(2):211. doi:10.1007/s12028-011-9605-9

8. Kim SM, Woo HG, Kim YJ, Kim BJ. Blood pressure management in stroke patients. J Neurocritical Care. 2020;13(2):69–79. doi:10.18700/jnc.200028

9. Lin QS, Ping-Chen, Lin YX, et al. Systolic Blood Pressure Variability is a Novel Risk Factor for Rebleeding in Acute Subarachnoid Hemorrhage: A Case–Control Study. Medicine (Baltimore). 2016;95(11):e3028. doi:10.1097/MD.0000000000003028

10. Svedung Wettervik T, Howells T, Enblad P, Lewén A. Intracranial pressure variability: relation to clinical outcome, intracranial pressure–volume index, cerebrovascular reactivity and blood pressure variability. J Clin Monit Comput. 2020;34(4):733–741. doi:10.1007/s10877-019-00387-9

11. Ajam K, Gold LS, Beck SS, Damon S, Phelps R, Rea TD. Reliability of the Cerebral Performance Category to classify neurological status among survivors of ventricular fibrillation arrest: a cohort study. Scand J Trauma Resusc Emerg Med. 2011;19(1):38. doi:10.1186/1757-7241-19-38

12. Grossestreuer AV, Abella BS, Sheak KR, et al. Inter-rater reliability of post-arrest cerebral performance category (CPC) scores. Resuscitation. 2016;109:21–24. doi:10.1016/j.resuscitation.2016.09.006

13. Quinn TJ, Dawson J, Walters MR, Lees KR. Exploring the Reliability of the Modified Rankin Scale. Stroke. 2009;40(3):762–766. doi:10.1161/STROKEAHA.108.522516

14. Nahm FS. Receiver operating characteristic curve: overview and practical use for clinicians. Korean J Anesthesiol. 2022;75(1):25–36. doi:10.4097/kja.21209

15. Svedung Wettervik T, Engquist H, Howells T, et al. Higher intracranial pressure variability is associated with lower cerebrovascular resistance in aneurysmal subarachnoid hemorrhage. J Clin Monit Comput. 2023;37(1):319–326. doi:10.1007/s10877-022-00894-2

16. Cai K, Zhang Y, Shen L, Ji Q, Xu T, Cao M. Characteristics of Blood Pressure Profiles After Endovascular Coiling as Predictors of Clinical Outcome in Poor-Grade Aneurysmal Subarachnoid Hemorrhage. World Neurosurg. 2017;104:459–466. doi:10.1016/j.wneu.2017.05.027

17. Germans MR, Coert BA, Vandertop WP, Verbaan D. Time intervals from subarachnoid hemorrhage to rebleed. J Neurol. 2014;261(7):1425–1431. doi:10.1007/s00415-014-7365-0

18. Larsen CC, Astrup J. Rebleeding After Aneurysmal Subarachnoid Hemorrhage: A Literature Review. World Neurosurg. 2013;79(2):307–312. doi:10.1016/j.wneu.2012.06.023

19. Gosmanova EO, Mikkelsen MK, Molnar MZ, et al. Association of Systolic Blood Pressure Variability With Mortality, Coronary Heart Disease, Stroke, and Renal Disease. J Am Coll Cardiol. 2016;68(13):1375–1386. doi:10.1016/j.jacc.2016.06.054

20. Parati G, Ochoa JE, Salvi P, Lombardi C, Bilo G. Prognostic Value of Blood Pressure Variability and Average Blood Pressure Levels in Patients With Hypertension and Diabetes. Diabetes Care. 2013;36(Supplement_2):S312–S324. doi:10.2337/dcS13-2043

21. Kirkness CJ, Burr RL, Mitchell PH. Intracranial and Blood Pressure Variability and Long-Term Outcome After Aneurysmal Sub-arachnoid Hemorrhage. Am J Crit Care. 2009;18(3):241–251. doi:10.4037/ajcc2009743

22. Hajian-Tilaki K. Receiver Operating Characteristic (ROC) Curve Analysis for Medical Diagnostic Test Evaluation. Casp J Intern Med. 2013;4(2):627–635.

23. Halligan S, Altman DG, Mallett S. Disadvantages of using the area under the receiver operating characteristic curve to assess imaging tests: A discussion and proposal for an alternative approach. Eur Radiol. 2015;25(4):932–939. doi:10.1007/s00330-014-3487-0

